# Identification of Early Risk Factors for Mortality in Pediatric Veno-Arterial Extra Corporeal Membrane Oxygenation: The Patient Matters

**DOI:** 10.1101/2024.10.17.24315712

**Authors:** Bennett Weinerman, Soon Bin Kwon, Tammam Alalqum, Daniel Nametz, Murad Megjhani, Eunice Clark, Caleb Varner, Eva W. Cheung, Soojin Park

**Affiliations:** Program for Hospital and Intensive Care Informatics, Department of Neurology, Columbia University Vagelos College of Physicians and Surgeons, New York, USA; Department of Pediatrics, Division of Critical Care & Hospital Medicine, Columbia University Vagelos College of Physicians and Surgeons and NewYork-Presbyterian Morgan Stanley Children’s Hospital, New York, USA; Department of Neurology, Columbia University Vagelos College of Physicians and Surgeons, New York, USA; Department of Nursing, NewYork-Presbyterian Morgan Stanley Children’s Hospital, New York, USA; Department of Perfusion, NewYork-Presbyterian Morgan Stanley Children’s Hospital, New York, USA; NewYork-Presbyterian Hospital, Columbia University Irving Medical Center, New York, USA; Department of Biomedical Informatics, Columba University, New York, USA

## Abstract

**Objective:** Pediatric Veno-Arterial Extra Corporeal Membrane Oxygenation (VA ECMO) is a life saving technology associated with high mortality. A successful VA ECMO course requires attention to multiple aspects of patient care, including ECMO and patient parameters. Early, potentially modifiable, risk factors associated with patient mortality should be analyzed and adjusted for when assessing VA ECMO risk profiles.

**Method:** Retrospective single center experience of pediatric patients requiring VA ECMO from January 2021 to October 2023. Laboratory and ECMO flow parameters were extracted from the patients record and analyzed. Risk factors were analyzed using a Cox proportion hazard regression

**Main Results:** There were 45 patients studied. Overall survival was 51%. Upon uncorrected analysis there were no significant differences between the patients who survived and those who died. Utilizing a Cox proportion hazard regression, platelet count, fibrinogen level and creatine level were significant risk factors within the first twenty-four hours of a patient’s ECMO course.

**Significance:** Although we did not find a significant difference among ECMO flow parameters in this study, this work highlights that granular ECMO flow data can be incorporated to risk analysis profiles and potential modeling in pediatric VA ECMO. This study demonstrated, that when controlling for ECMO flow parameters, kidney dysfunction and clotting regulation remain key risk factors for pediatric VA ECMO mortality.

## Introduction

Extra Corporeal Membrane Oxygenation (ECMO) was first introduced as a viable medical therapy in the 1970s in the setting of severe acute respiratory distress syndrome (ARDS) (1). Since that time, ECMO has undergone major refinement, leading to increased utilization and improved outcomes. Veno-Arterial ECMO (VA-ECMO) treats patients who are often on the brink of viable physiology, and thus are at extremely high risk of mortality. VA ECMO is increasingly implemented in pediatric patients, with mortality improving over the past decades with current literature citing a mortality around 40-60% (2-7).

Decisions regarding goals of care are often clouded by the inability of the clinical team to provide accurate prognostic information. Accurate risk factors for poor outcomes allow the clinical team to have more informed conversations with care givers. Studies have tried to predict which patients will have better outcomes after implementation of ECMO support. Several adult studies have leveraged machine learning algorithms (8-10); however, these models largely rely on registry data(11). Many pediatric ECMO research studies have found that age, gender, the development of renal or hepatic dysfunction were associated with mortality. Other groups who have included more diverse pediatric patient populations have found that lactate, pH (both before and after cannulation), as well as, renal dysfunction, including use of continuous renal replacement, and active cannulation during cardiopulmonary resuscitation (eCPR) are associated with mortality(4-6, 12, 13). Other variables, such as single-ventricle physiology, location of patient cannulation and length of ECMO run are often, but not always, sited as significant risk factors(6, 12). Novel non-invasive metrics, like echocardiographic studies have also been associated with pediatric ECMO survival (3). Notably, these studies do not rigorously analyze the ECMO derived flow parameters for each patient.

A successful VA-ECMO run requires attention to detail from multifaceted team members. There are multiple technical as well as patient and ECMO physiologic variables that must be continuously adjusted for optimal outcomes. This retrospective observational cohort study investigated routinely collected laboratory data, as well as ECMO flow data within the first 24 hours of the ECMO run to determine whether to elucidate early risk factors for patient mortality once on VA ECMO. Our aim was to focus on early metrics with a goal of minimizing confounding variables.

## Methods

### Study Population

This is a retrospective observational cohort study at a single academic institution with both a neonatal and pediatric cardiac intensive care unit. Pediatric patients who were treated with VA ECMO between January 2021 and October 2023 were included. Location of cannulation included operating room, PICU, Neonatal Intensive Care Unit (NICU) and in other hospital locations (i.e. cardiac catheterization lab) were recorded. The underlying indication for the patient’s decompensation, necessitating the need for VA ECMO, was extracted from their medical records based on progress notes. Patients were excluded from analysis based on the following criteria: (1) ECMO run lasting less than 24 hours; (2) multiple ECMO runs; (3) single-ventricle anatomy; (4) missing data due to cannulation location or incomplete data. The Columbia University IRB committee approved this research study (Protocol # AAAU5398). Research was conducted in accordance with the principles embodied in the Declaration of Helsinki and in accordance with local statutory requirements. This study was a non-treatment, retrospective, observational review of physiologic and ECMO data that was obtained as part of routine standard of care, and the IRB approved a waiver of informed consent.

### ECMO Circuit

Our institution performs 40-50 VA ECMO runs per year. ECMO cannulation was performed by a member of the Pediatric Cardiothoracic Surgery or Pediatric General Surgery Team. Central or peripheral cannulation was at the discretion of the surgical team based on the timing of the most recent sternotomy. ECMO support was provided by the Cardiohelp ECMO system from Getinge (Göteborg, Sweden), with either a 5.0 oxygenator for patients <19kg or the 7.0 oxygenator for children >19kg. A perfusionist adjusted the ECMO settings in consultation with the clinical team. Our hospital does not have a protocol or pathway for weaning ECMO support. Anti-coagulation was adjusted per institution protocol based on bleeding and non-bleeding conditions.

### Data Collection

Physiologic data was collected by the bedside monitor as part of routine standard of care. ECMO flow data was retrospectively extracted from the Spectrum module (Gloucester, UK) at a sampling frequency of 0.016Hz or 1 sample per minute. The first 24-hours of data after ECMO cannulation were included for analysis. The start of the ECMO run was defined as the time that ECMO parameters were output from the Spectrum data file. This date and time stamp was corroborated with our internal ECMO database to ensure validity. ECMO Flow data consisted of Arterial Flow, Delta Pressure, Fractional Delivery of Oxygen (FDO_2_), Arterial Pressure, Venous Pressure, Cardiac Index (CI), Revolutions Per Minute (RPM), SaO_2_, SpO_2_, SvO_2_, and Sweep. Laboratory data included Hemoglobin, Hematocrit, Platelet Count, Arterial pH, Arterial partial pressure of carbon dioxide (paCO_2_), arterial bicarbonate (HCO_3_), Creatinine, Fibrinogen, Aspartate Aminotransferase (AST), Alanine Aminotransferase (ALT), and plasma free hemoglobin.

### Statistical Analysis

All data analyses were performed using MATLAB 2020a (MathWorks, Massachusetts). For each patient, the average value was calculated over the 24-hour period of the ECMO run for each variable. For each group (i.e. Deceased or Survived), an average was calculated. Students’ *t*-test with a significance level of ≤0.05 was performed to compare the difference between the two groups. For demographic and baseline characteristics between the two groups, continuous variables were compared using a non-paired Student’s *t*-test, assuming heterogenous variance. Categorical variables were compared using Fisher’s exact test, or Chi-squared test when appropriate with a significance level of ≤0.05. Cox proportion hazard regression was performed to identify risk factors for 180-day mortality. Among the variables with high correlation, only one variable was included to reduce bias introduced to the analysis. For example, for AST and ALT, only AST was used. Similarly, only hematocrit was incorporated into the model from the choice of hemoglobin or hematocrit. While it is recognized that pH, pCO_2_ and sweep are all interconnected, given their clinical significance, all three variables were included in the model. Similarly, RPM and CI were both included in the regression given their clinical implications.

## Results

There were 106 patients who were cannulated to VA-ECMO between January 2021 and October 2023. Nine patients had multiple ECMO runs, and the 22 patients who had single ventricle anatomy were removed from analysis. Seven, nine and fourteen patients had an ECMO run <24 hours, were cannulated at an outside hospital and subsequently transferred to our institution or had missing data respectively, and were removed from analysis. Of the remaining 45 patients 23 (51%) survived and 22 (49%) died. Twelve (27%) patients were cannulated in the setting of respiratory indications, twenty-three (51%) were cannulated for primarily cardiac indications and ten (22%) were cannulated in the setting of eCPR (**Table 1**).

**Table 1:** Demographic information for patients analyzed on Veno-Arterial Extracorporeal Membrane Oxygenation. (PICU = Pediatric Intensive Care Unit, NICU = Neonatal Intensive Care Unit, OR = Operating Room, Cath Lab = Cardiac Catheterization Laboratory, ECMO = Extracorporeal Membrane Oxygenation, pHTN = Pulmonary Hypertension, OHT = Orthotopic Heart Transplant, CDH = Congenital Diaphragmatic Hernia, DCM = Dilated Cardiomyopathy, RCM = Restrictive Cardiomyopathy, HLH = Hemophagocytic Lymphohistiocytosis)

|  | **Total**<br>**(45)** | **Survived**<br>**(23)** | **Deceased**<br>**(22)** | **p -**<br>**value** |
| --- | --- | --- | --- | --- |
| Age at cannulation<br>(months) | 54.0 | 48.81 | 59.47 | 0.66 |
| Gender (F:M) | 23:22 | 11:12 | 12:10 | 0.77 |
| Weight (kg) | 16.9 | 15.2 | 18.8 | 0.57 |
| Cannulation |  |  |  | 0.75 |
| Central | 13 | 6 | 7 |  |
| Peripheral | 32 | 17 | 15 |  |
| Cannulation Location |  |  |  | 1.00 |
| PICU | 23 | 12 | 11 |  |
| NICU | 14 | 7 | 7 |  |
| OR | 6 | 3 | 3 |  |
| Cath Lab | 2 | 1 | 1 |  |
| Indication |  |  |  | 0.41 |
| Cardiac | 23 | 14 | 9 |  |
| Respiratory | 12 | 5 | 7 |  |
| eCPR | 10 | 4 | 6 |  |
| Length of ECMO run<br>(hours) | 182.79 | 178.18 | 187.60 | 0.85 |
| Diagnosis (Top 10) |  |  |  |  |
| pHTN | 8 | 6 | 2 |  |
| Valvular Disease | 5 | 3 | 2 |  |
| OHT Complication | 4 | 1 | 3 |  |
| CDH | 4 | 3 | 1 |  |
| DCM | 3 | 3 | 0 |  |
| RCM | 3 | 2 | 1 |  |
| Infection | 2 | 0 | 2 |  |
| Tetralogy of Fallot | 2 | 2 | 0 |  |
| HLH | 2 | 2 | 0 |  |

The study included 23 (51%) females. The patients’ ages ranged from birth (0 days) to 22 years old with an average of 4.5 years old (54 months). The average patient’s weight prior to cannulation was 16.9kg. The average ECMO run was 182.78 hours. There was no significant difference in patient demographic data comparing those who survived, versus those who died (**Table 1**).

Twenty-three patients were cannulated while in the PICU (51.1%). Fourteen patients were cannulated in the NICU (31.1%). Six patients (13.3%) were cannulated in the operating room and two patients (4.4%) were cannulated in the cardiac catheterization lab.

The most common diagnoses were pulmonary hypertension (8 patients), Valvular disease (5 patients), Heart transplant complications (4 patients), congenital diaphragmatic hernia (4 patients), dilated cardiomyopathy (3 patients), restrictive cardiomyopathy (3 patients), infection (2 patients), Tetralogy of Fallot (2 patients) and Hypoplastic Left Heart (2 patients). For a complete breakdown of the entire cohort please see ***supplementary table 1***.

Lab values and ECMO flow data were averaged over the first 24-hour period of the ECMO run per patient and then averaged for each cohort (survived or deceased). For the laboratory values within the first 24 hours of the ECMO run, the average ALT (Units/Liter) was 543.7 in those who survived compared to 455.9 in those who died (*p* = 0.77). The average fibrinogen (mg/dL) was 192.1 versus 151.11 (*p* = 0.053) between the survivors and the deceased respectively. The average hematocrit (%) among the survivors was 32.0 versus 31.2 in the deceased patients (*p*=0.56). The average platelet count (×10^3^/µL) was 111.5 and 104.4 (*p* = 0.68) in the patients who survived versus those who died. Among survivors the average creatinine (mg/dL) was 0.67 versus 1.01 (*p* = 0.067) in those who died. The average pH and paCO_2_ (mmHg) was 7.34 and 41.7 in the survivors compared to 7.36 and 42.5 in those who died respectively, with corresponding *p*-values of 0.56 and 0.64. The average lactate level (mmoL/L) was 3.68 in survivors and 4.16 (*p* = 0.66) in those patients who died (**Table 2**)

**Table 2:** Averaged values and standard deviation for each patient cohort, survived versus deceased. (paCO_2_ = arterial partial pressure of carbon dioxide, ALT = alanine aminotransferase, CI = Cardiac Index, RPM = Revolutions per Minute, FDO2 = Fraction of Delivered Oxygen)

| **Laboratory and ECMO flow Data**<br>**Mean and Standard Deviation** |  |  |  |
| --- | --- | --- | --- |
|  | Survived<br>(n=23) | Deceased<br>(n=22) | p-value |
| Arterial pH | 7.34 (0.09) | 7.36 (0.06) | 0.56 |
| paCO<sub>2</sub> (mmHg) | 41.73 (5.86) | 42.53 (5.42) | 0.64 |
| Lactate (mmol/L) | 3.68 (3.5) | 4.16 (3.8) | 0.66 |
| Hematocrit (%) | 32.01 (4.9) | 31.22 (4.2) | 0.56 |
| Platelet (×10<sup>3</sup>μL) | 111.5 (60.5) | 104.4 (55.3) | 0.68 |
| Fibrinogen (mg/dL) | 192.08 (79.8) | 151.11 (55.2) | 0.053 |
| Creatinine (mg/dL) | 0.67 (0.4) | 1.01 (0.8) | 0.067 |
| ALT (U/L) | 543.7 (1210.4) | 455.9 (659.0) | 0.77 |
| CI (L/min/m<sup>2</sup>) | 1.94 (0.4) | 2.13 (0.3) | 0.073 |
| RPM | 2708.18 (317.2) | 2854.12 (443.6) | 0.21 |
| Sweep (L/min) | 0.96 (1.7) | 1.04 (1.7) | 0.87 |
| FDO<sub>2</sub> | 59.69 (21.1) | 60.62 (19.8) | 0.88 |

For the ECMO flow values the average CI (L/min/m^2^) was 1.94 in survivors versus 2.13 in those who died (*p* = 0.073). The corresponding average RPMs were 2708.2 compared to 2854.1 (*p* = 0.21) in those patients who survived and those who died. The average ECMO sweep was 0.96 in the patients who survived compared to 1.0 (*p* = 0.87) in those who lived. The FDO2 of the ECMO circuit averaged 59.7 for patients who survived compared to 60.6 (*p* = 088) for patients who died. (**Figure 1**).

**Figure 1:**
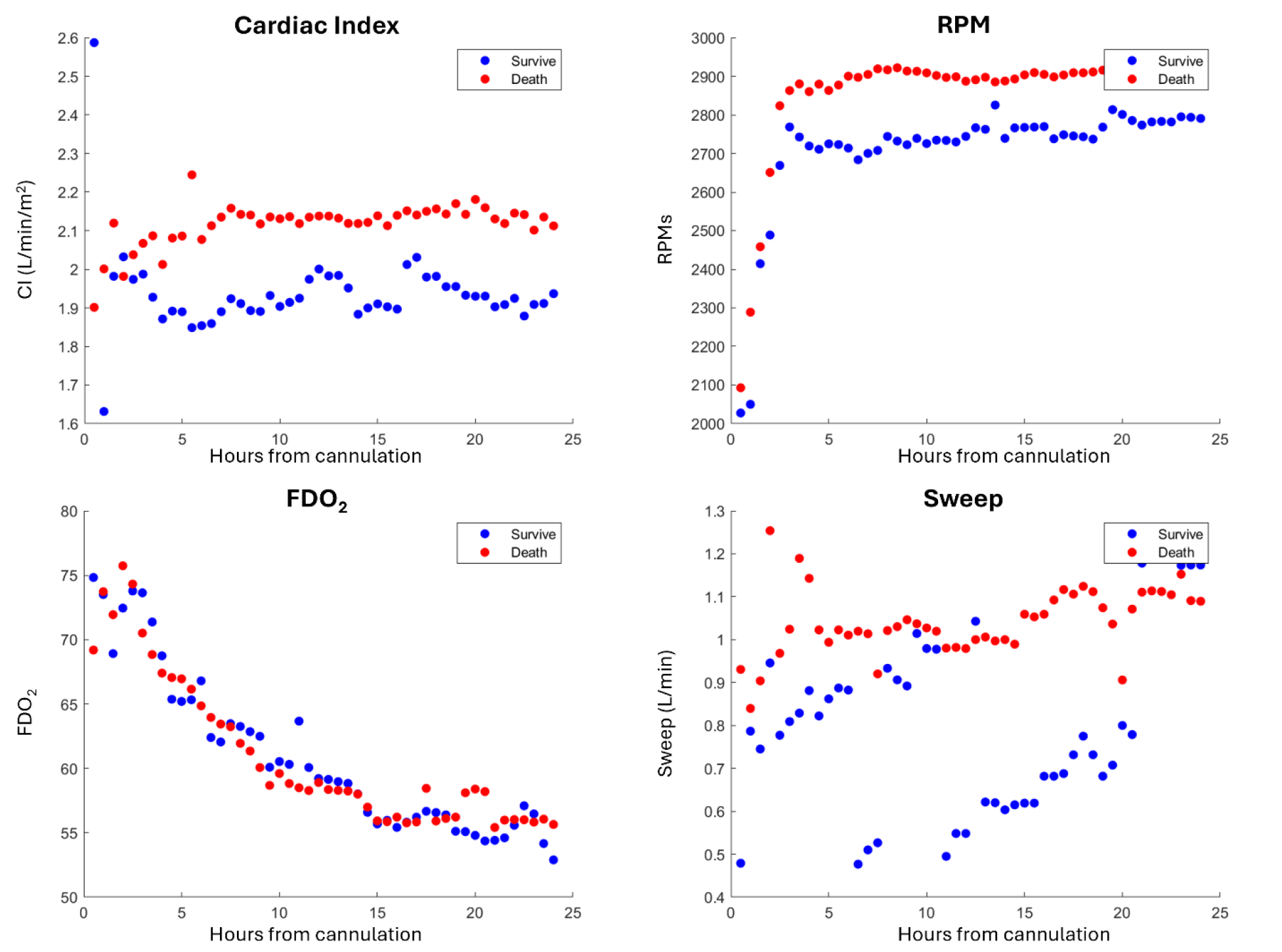
Averaged ECMO parameters every hour from time of ECMO cannulation, grouped on those who survived versus those who died. CI = Cardiac Index, RPM = Revolutions per Minute, FDO2 = Fraction of Delivered Oxygen.

A Cox regression was run using the same variables above. The 180-day survival analysis is shown in **Table 3**. After running the Cox survival analysis only three values were significant. For fibrinogen, the Cox coefficient was -0.015, with a hazard ratio of 0.99 and a *p* value of 0.004. The corresponding 95^th^ percentile hazard ratio confidence interval ranged from 0.98 to 1.00. For creatinine the Cox coefficient was 1.14 with a hazard ratio of 3.12 and a *p* value of 0.022. The 95^th^ percentile hazard ratio confidence interval ranged from 1.18 to 8.27 for Creatinine. The Cox coefficient for Platelet count was 0.011, with a hazard ratio of 1.01 and a *p* value of 0.04. The 95^th^ percentile hazard ratio confidence interval ranged from 1.00 to 1.02.

**Table 3:** Cox Hazard Coefficient and Hazard ratio for each variable analyzed between the two cohorts, survived versus deceased. (paCO_2_ = arterial partial pressure of carbon dioxide, ALT = alanine aminotransferase, CI = Cardiac Index, RPM = Revolutions per Minute, FDO2 = Fraction of Delivered Oxygen)

|  | Coefficient | Hazard Ratio | p-value | Hazard Ratio<br>95% CI |  |
| --- | --- | --- | --- | --- | --- |
| pH | 4.31 | 74.1 | 0.33 | 0.013 | 430761 |
| paCO<sub>2</sub> (mmHg) | 0.027 | 1.03 | 0.50 | 0.95 | 1.11 |
| Lactate (mmol/L) | 0.14 | 1.14 | 0.15 | 0.95 | 1.38 |
| Hematocrit (%) | - 0.044 | 0.96 | 0.49 | 0.85 | 1.08 |
| Platelet (×10<sup>3</sup>μL) | 0.011 | 1.01 | 0.04 | 1.00 | 1.02 |
| Fibrinogen (mg/dL) | - 0.015 | 0.99 | 0.004 | 0.98 | 1.00 |
| Creatinine (mg/dL) | 1.14 | 3.12 | 0.02 | 1.18 | 8.27 |
| ALT (U/L) | -0.0005 | 1.00 | 0.073 | 1.00 | 1.00 |
| CI (L/min/m<sup>2</sup>) | 0.44 | 1.55 | 0.60 | 0.30 | 8.01 |
| RPM | 0.0007 | 1.00 | 0.47 | 1.00 | 1.00 |
| Sweep (L/min) | 0.017 | 1.02 | 0.95 | 0.58 | 1.79 |
| FDO<sub>2</sub> | - 0.012 | 0.99 | 0.49 | 0.96 | 1.02 |

## Discussion

Among patients requiring a singular run of VA ECMO we found a survival rate of 51%. This survival rate is consistent with previously cited pediatric studies and congruent with data published by the ELSO network (14-17). Our analysis was comprised of pediatric patients recovering after cardiac surgery as well as patients admitted for other medical indications who subsequently required VA ECMO, including those patients who required eCPR.

Within the first 24 hours of VA ECMO cannulation, we did not find any laboratory or ECMO parameter that were significantly different among those patients who survived versus those who died. Upon using a 180-day Cox regression, we found that fibrinogen (*p* = 0.004), creatinine (*p* = 0.02) and platelet count (*p* = 0.04) were significant risk factors associated with mortality within the first 24 hours of a patients ECMO run.

Renal dysfunction and/or requiring continuous renal replacement therapy (CRRT) while on ECMO has been reported as a significant risk factor for mortality (16, 18). Additionally, decreased urine output, likely a measure of both end organ perfusion and ongoing nephrogenic insult has been cited as a risk factor for pediatric VA ECMO mortality (15). Authors have gone on to stratify the degree of renal dysfunction, based on the Kidney Disease Improving Global Outcome (KDIGO) definition, and its impact on in-hospital mortality for patients requiring VA ECMO (17). Our findings suggest that the risk associated with kidney disease is evident within the first 24 hours of a patients ECMO course; even while controlling for the impact of other ECMO parameters.

Additionally, a patient’s hematologic risk profile has been associated with mortality (19). We found that platelet and fibrinogen levels are risk factors for patient survival. Interestingly, our data suggests that higher fibrinogen levels may be slightly protective, evident by a hazard ratio less than 1. Conversely, a higher platelet count was associated with a marginal increased risk of mortality. It is important to note the complexity of the clotting cascade, and how both fibrinogen and platelet are impacted by systemic inflammation, bleeding and the ECMO circuit itself. Our findings further highlight how critical appropriate anti-coagulation is when managing pediatric VA ECMO (20-22).

We hoped to eliminate some confounding factors by focusing on the first 24 hours of a patient’s ECMO course, where ideally the ECMO oxygenator and tubing is relatively devoid of significant fibrin deposition (19). However, we cannot account for the microscopic changes that occur at the blood-oxygenator membrane(23-25), or the inflammatory changes that occur due to the presence of the ECMO cannulations themselves (26). Unfortunately, within the first 24 hours of a patients ECMO run, we did not have enough data to comment on the role of plasma free hemoglobin, or other clotting factors. Additionally, our institution only uses centrifugal ECMO circuits, and the implication on roller versus centrifugal systems is not addressed in our study.

Laboratory evidence of poor organ perfusion is frequently cited as being risk factors for patient mortality while on ECMO. In our regression, we did not find hepatic enzymes (ALT), lactate or pH to be significant (15, 17, 27). As we only looked at data within the first 24 hours, it is likely that some of these markers would have become significant given a longer sampling window. We were unable to include the partial pressure of oxygen (paO_2_) as a laboratory parameter, given the uncertainty surrounding this lab value. Given the retrospective nature of this study we were not able to determine if the paO_2_ values were taken from the patient or the ECMO circuit, and thus were not analyzed.

To our knowledge this is the first pediatric study to examine relatively high frequency ECMO variables in conjunction with patient laboratory markers. Given the complexity of ECMO, we believe it is imperative for retrospective studies to incorporate the impact that the ECMO circuit and ECMO parameters have on the patient’s survival; however, in our study, ECMO circuit parameters were not significant risk factors within the first 24 hours. It seems clear to the practicing clinician, that the odds of survival are much different for a child requiring a CI of 3.5 L/min/m^2^ versus 1.5 L/min/m^2^, or a sweep of 10 L/min versus a sweep of 0.3 L/min, yet these variables were not significant during the first 24 hours of a patient’s ECMO course. This study further demonstrates that ECMO flow parameters can be captured and should be controlled for, in any risk analysis. Additionally, we selected laboratory and ECMO parameters that are frequently cited as being risk factors for patient outcomes. We decided to focus on the first 24 hours of a patient’s ECMO run, hoping to elucidate early modifiable risk factors that may change a patients’ outcome, while limiting other confounding variables.

This was a retrospective study of pediatric patients requiring VA ECMO and thus has limitations. We included patients who were cannulated both centrally and peripherally, thus this study includes a heterogeneous population. Furthermore, our institution does not have strict criteria as to which patients get cannulated onto ECMO, thus there is even more heterogeneity based on the anesthesiologist/intensivist and the surgeon involved in the cannulation. Our sample size is relatively large for a single center; however, more patients would better elucidate the significance of each risk factor. Additionally, we did not incorporate physiologic parameters in this analysis (i.e. heart rate, mean arterial pressure etc.), which we hope to do in subsequent studies.

## Conclusion

Over a 3-year period we analyzed 45 pediatric patients who were cannulated onto VA-ECMO with a 51% survival rate. While there were no overt significant differences between the two groups, using a Cox regression including key ECMO parameters, we found that creatinine, fibrinogen and platelet count are key risk factors for patient mortality within the first 24 hours of a patient’s ECMO run. Our data provide evidence that maximal effort should be spent optimizing the patient, as none of the ECMO parameters were significant risk factors for patient mortality.

The use of mechanical circulatory support is a complex and nuanced science that has provided lifesaving therapy to a heterogenous population. For accurate and impactful prediction, it is necessary to capture as many aspects of these patients’ care as possible, including ECMO support parameters. We hope that with more multifaceted, granular clinical data, we will be better able to arm the clinical team with accurate and predictive information.

## Data Availability

Data for this study cannot be readily de-identified and thus data will not be shared.

## Supplemental Table

**Supplementary Table 1:** Diagnoses of patients included in the analysis.

| Diagnosis associated with ECMO Cannulation | Total Count | Survived | Deceased |
| --- | --- | --- | --- |
| Pulmonary Hypertension | 8 | 2 | 6 |
| Valvular Disease | 5 | 3 | 2 |
| Congenital Diaphragmatic Hernia | 4 | 3 | 1 |
| Heart Transplant Complication | 4 | 1 | 3 |
| Dilated Cardiomyopathy | 3 | 3 | 0 |
| Restrictive Cardiomyopathy | 3 | 2 | 1 |
| Tetralogy of Fallot | 2 | 2 | 0 |
| Infection | 2 | 0 | 2 |
| Hemophagocytic lymphohistiocytosis | 2 | 2 | 0 |
| Cardiac Mass | 1 | 1 | 0 |
| Heart Failure | 1 | 0 | 1 |
| Alveolar Capillary Dysplasia | 1 | 0 | 1 |
| Double Aortic Arch | 1 | 1 | 0 |
| Double Outlet Right Ventricle with Transposition of the Great Arteries | 1 | 1 | 0 |
| d-Transposition of the Great Arteries | 1 | 1 | 0 |
| Hypertrophic Cardiomyopathy | 1 | 1 | 0 |
| Atrio-Ventricular Canal | 1 | 0 | 1 |
| Partial Anomalous Pulmonary Venous Return | 1 | 0 | 1 |
| Asthma | 1 | 1 | 0 |

